# Defining the Clinical Validity of Genes Reported to Cause Pulmonary Arterial Hypertension

**DOI:** 10.1101/2022.09.02.22279461

**Authors:** Carrie L. Welch, Micheala A. Aldred, Srimmitha Balachandar, Dennis Dooijes, Christina A. Eichstaedt, Stefan Gräf, Arjan C. Houweling, Rajiv D. Machado, Divya Pandya, Matina Prapa, Memoona Shaukat, Laura Southgate, Jair Tenorio-Castano, the ClinGen PH VCEP, Wendy K. Chung, the International Consortium for Genetic Studies in Pulmonary Arterial Hypertension (PAH-ICON) at the Pulmonary Vascular Research Institute (PVRI)

## Abstract

**BACKGROUND:** Pulmonary arterial hypertension (PAH) is a rare, progressive vasculopathy with significant cardiopulmonary morbidity and mortality. The disease is caused by both genetic and environmental factors, with genetic variants in at least 27 genes displaying putative evidence for disease causality. Genetic testing is currently recommended for adults diagnosed with heritable or idiopathic PAH, and all children diagnosed with PAH. However, testing panels vary in the number and list of genes included, and exome/genome sequencing data may reveal variants in genes with varying levels of evidence for a relationship with PAH.

**METHODS:** An international panel of clinical and scientific experts in PAH was formed to perform an evidence-based review of heritable and idiopathic PAH gene-disease relationships. The panel performed literature searches and applied a semi-quantitative scoring system developed by the NIH Clinical Genome Resource to classify the relative strength of PAH gene-disease relationships based on genetic and experimental evidence.

**RESULTS:** Of twenty-seven genes curated, twelve genes (*BMPR2, ACVRL1, ATP13A3, CAV1, EIF2AK4, ENG, GDF2, KCNK3, KDR, SMAD9, SOX17*, and *TBX4*) were classified as having definitive evidence for causal effects of variants. Three genes, *ABCC8, GGCX*, and *TET2*, were classified as having moderate evidence. Six genes (*AQP1, BMP10, FBLN2, KLF2, KLK1*, and *PDGFD*) were classified as having limited evidence, and TOPBP1 was classified as having no known PAH relationship. Some of the recently identified genes with moderate or limited evidence may move to a higher classification as new evidence emerges. Five genes (*BMPR1A, BMPR1B, NOTCH3, SMAD1*, and *SMAD4*) were disputed due to a paucity of genetic evidence over time.

**CONCLUSIONS:** Evidence-based classification of PAH gene-disease relationships indicates that twelve genes have definitive evidence for causal effects of variants. We recommend that genetic testing panels include all genes with definitive evidence and that caution be taken in the interpretation of variants identified in genes with moderate or limited evidence. Genes with no known evidence for PAH or disputed genes should not be included in testing panels.

**Clinical Perspective:** *What is New?:*

- Evidence-based PAH gene curation was performed using the NIH Clinical Genome Resource model.
- Heritable and idiopathic PAH are caused by pathogenic variants in a diverse set of genes, including genes in the TGFβ/BMP pathway, channelopathy genes, cell metabolism genes, growth factors and transcription factors.
- Four previously reported TGF-β/BMP pathway genes are disputed for a PAH gene-disease relationship.

*What Are the Clinical Implications?:*

- All genes with definitive evidence for a PAH gene-disease relationship are strongly recommended to be included in genetic testing panels.
- Caution should be taken in clinical interpretation for genes with less than definitive or strong evidence and disputed genes or genes with no known genetic evidence for PAH should not be included in genetic testing panels.
- For undiagnosed cases, genetic reanalysis is recommended over time as new evidence for PAH gene-disease relationship is evaluated.

## INTRODUCTION

Pulmonary arterial hypertension (PAH) is a rare, progressive, and often lethal disease characterized by distinctive changes in pulmonary arteries that leads to increased pulmonary vascular resistance, right ventricular (RV) hypertrophy, and, ultimately, right heart failure (1-3). PAH can occur in families or sporadically, and either idiopathically or associated with other diseases such as congenital heart disease, autoimmune connective tissue diseases, portopulmonary disease, or associated with exposure to certain medications/toxins. PAH may be caused by genetic, epigenetic, and environmental factors, as well as gene-environment interactions wherein genetic contributions to disease risk are modified by environmental exposures. *BMPR2* (bone morphogenetic protein receptor 2) is the most frequently identified causal gene for heritable PAH (HPAH), but at least twenty-six additional genes have been implicated in idiopathic PAH (IPAH) and some HPAH cases. Moreover, the use of exome and genome sequencing approaches has increased the rate of new candidate risk gene reporting. Independent validation of PAH gene-disease relationships is critical to avoid over-interpretation of genetic findings and reporting of large numbers of variants of uncertain significance (VUS) which can result in patient anxiety and influence life choices. Variable levels of evidence for each PAH gene-disease relationship complicate the clinical interpretation of genetic testing results and the prioritization of research strategies in the field. Currently, education about the option of genetic testing is strongly recommended for H/IPAH adults and all pediatric PAH patients. Thus, systematic review of the strength of evidence for PAH gene-disease relationships is needed.

We assembled an international panel of clinical geneticists, molecular genetic scientists, and computational genomic scientists with relevant expertise to systematically classify genes based on the strength of evidence for PAH gene-disease relationships. We applied the National Institutes of Health Clinical Genome Resource (ClinGen) (4) framework for semiquantitative classification of gene-disease relationships (5), under the auspices of the ClinGen Cardiovascular Domain Working Group. Here, we report the results of evidence-based gene curation for twenty-seven genes implicated in PAH.

## METHODS

### Study design and criteria

We assembled a ClinGen pulmonary hypertension gene curation expert panel (PH GCEP) (https://clinicalgenome.org/affiliation/40071/) consisting of sixteen members from eight institutions and representing six countries. An overview of the gene classification process and scoring criteria is provided in Figure 1. The scope of work included genes implicated in isolated H/IPAH: *BMPR2, ABCC8, AQP1, ATP13A3, BMP10, BMPR1A, BMPR1B, CAV1, EIF2AK4, FBLN2, GDF2, GGCX, KCNK3, KDR, KLF2, KLK1, NOTCH3, PDGFD, SMAD1, SMAD4, SMAD9, SOX17, TET2*, and *TOPBP1*. We also curated three genes implicated in syndromic forms of PAH: *ACVRL1* (PAH associated with hereditary hemorrhagic telangiectasia, HHT), *ENG* (PAH-HHT), and *TBX4* (small patella or *TBX4* syndrome) (6). Cases in which PAH was associated with other diseases and persistent pulmonary hypertension of the newborn were excluded from the curations. Genes were assigned to expert panel members taking into consideration conflicts of interest. Any member directly involved in gene discovery for a specific gene-PAH relationship(s), could not be assigned to lead the curation effort for that gene(s) or influence the group discussion of the final classification. Extensive literature searches were performed for each gene-disease relationship to identify relevant genetic and experimental evidence. A total of 168 peer-reviewed reports were evaluated for evidence. Data from several moderate-to large-sized cohorts were included in multiple gene curations and the cohorts are described here for brevity. The UK NIHR Bioresource – Rare Diseases Study for PAH is a case-control study including 1,038 unrelated, primarily European adult IPAH patients with genome sequencing data (herein, UK NIHR PAH cohort) (7-9). The National Biological Sample and Data Repository for PAH study is a US case-control study including 2,572 unrelated pediatric and adult PAH cases (43% IPAH, 48% PAH associated with other diseases, 4% HPAH and 5% other PAH) of mixed ancestry and with exome sequencing data (herein, PAH Biobank) (8-11). Wang *et al* is a cohort of 331 Han Chinese IPAH cases (Han Chinese IPAH cohort) with exome or genome sequencing (12). The Spanish Registry includes 300 samples from individuals with primary and associated forms of PAH for which panel or exome sequencing was performed (13-16).

**Figure 1.**
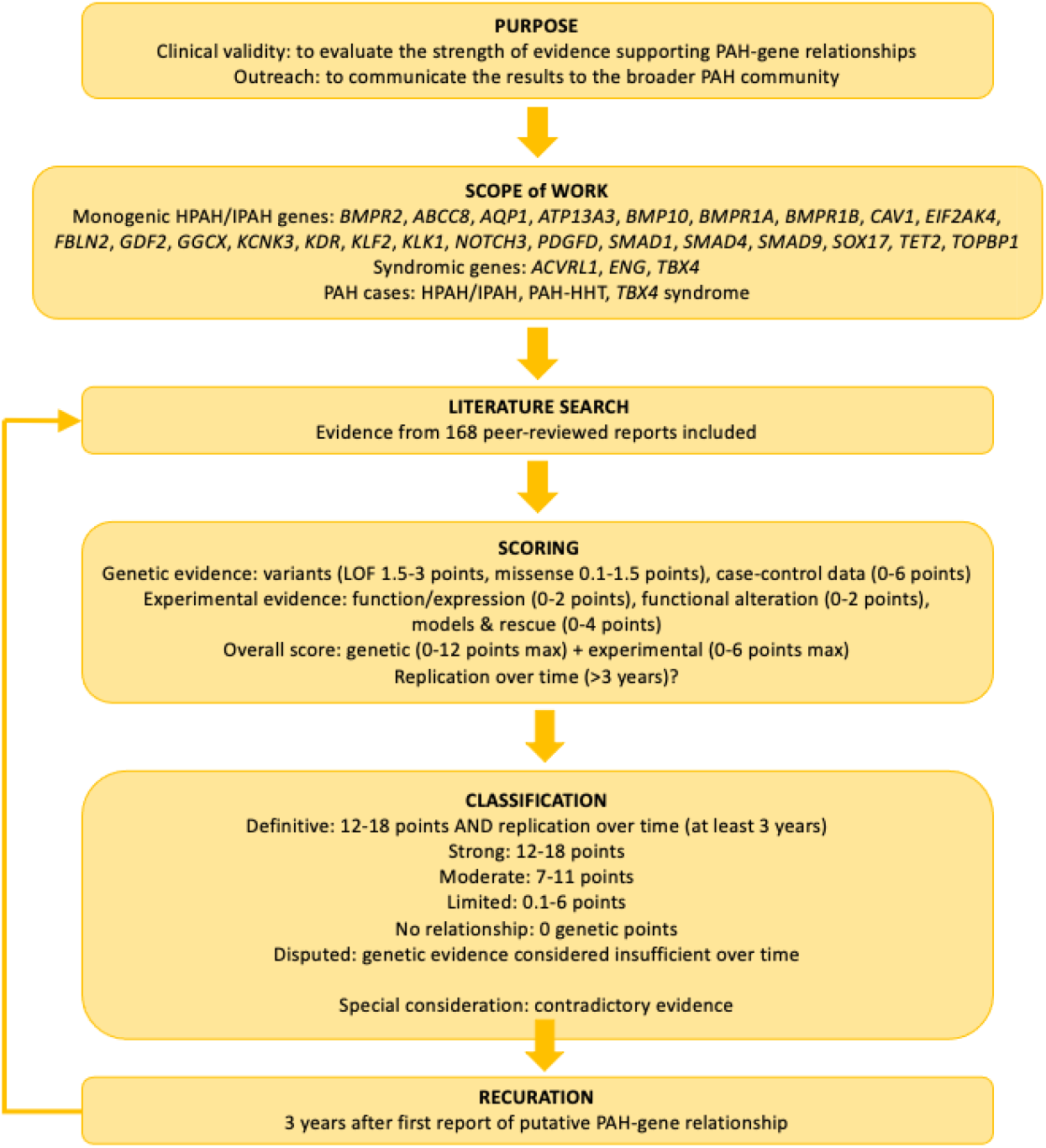
Gene curation flow chart.

### Genetic and functional evaluations of gene-disease relationships

Lead curators scored genetic and experimental evidence according to the updated ClinGen framework (Standard Operating Protocol v9.0). The predominant type of genetic evidence scored was case-level variant data. The threshold for inclusion was minor allele frequency less than 1/10,000 (gnomAD all, v2.1.1 controls, or relevant genetic ancestry) and variant type likely gene-disrupting (LGD; nonsense, frameshift, consensus splice variant) or missense with *in silico* predictions of deleteriousness ((CADD score ≥ 20 (17) or REVEL score with gene-specific thresholds (10)). Case-level evidence scores were weighted based on variant type, available functional data, and *de novo* inheritance. Family segregation and case-control association analyses were rarely scored due to incomplete penetrance of PAH risk alleles and PAH being a rare disease, respectively. Experimental evidence included expression in PAH relevant tissues/cells; known function including signaling pathways (for example, the BMP phosphorylation cascade), cell proliferation, apoptosis, and others; functional alteration in cells with patient variants; and PAH-relevant animal models and rescue. Scores were summed as defined by SOP v9.0 and individual PAH gene-disease relationships were classified as strong, moderate, limited, no known relationship, or disputed due to a lack of genetic evidence over time. Definitive classifications were assigned to genes with strong evidence for causality (12-18 points) plus independent evaluation of data over a period of at least 3 years post-discovery without contradictory evidence. Provisional gene curations were presented to the full PH GCEP via monthly video calls. Genes curated before the 3-year post-discovery timepoint underwent recuration at the 3-year timepoint. Final classifications were voted on by the group and curations were published to ClinGen’s website. All PH GCEP gene curations are publicly available on the ClinGen PH GCEP webpage (https://search.clinicalgenome.org/kb/affiliate/10071) or in collaboration with the ClinGen Hemostasis/Thrombosis GCEP https://search.clinicalgenome.org/kb/affiliate/10028) where indicated.

## RESULTS

### 1. Strength of evidence for genes implicated in isolated H/IPAH

Of twenty-four genes curated for isolated H/IPAH, nine of the gene-disease relationships were classified as definitive (*ATP13A3, BMPR2, CAV1, EIF2AK4, GDF2, KCNK3, KDR, SMAD9, SOX17*), three as moderate (*ABCC8, GGCX, TET2*), and six had limited evidence at the time of curation (*AQP1, BMP10, FBLN2, KLF2, KLK1, PDGFD*). One gene was classified as having no known relationship (*TOPBP1*) and five genes were classified as disputed (*BMP1A, BMPR1B, NOTCH3, SMAD1, SMAD4*). We note that *EIF2AK4* was curated based on pulmonary veno-occlusive disease (PVOD)/pulmonary capillary hemangiomatosis (PCH) which is often misdiagnosed as IPAH. Tabular and graphical summaries of these curations are provided in Table 1 and Figure 2.

**Table 1.**
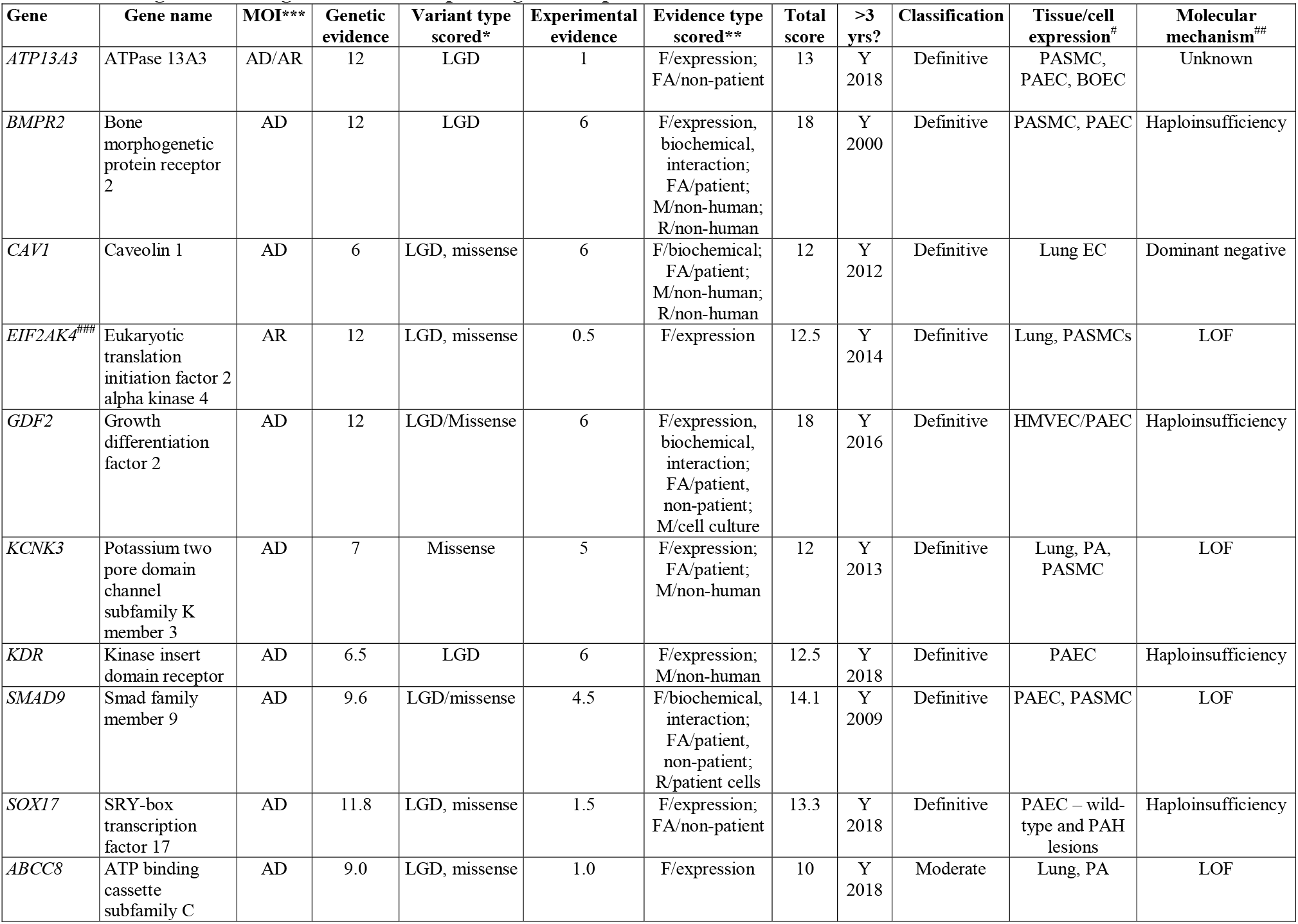
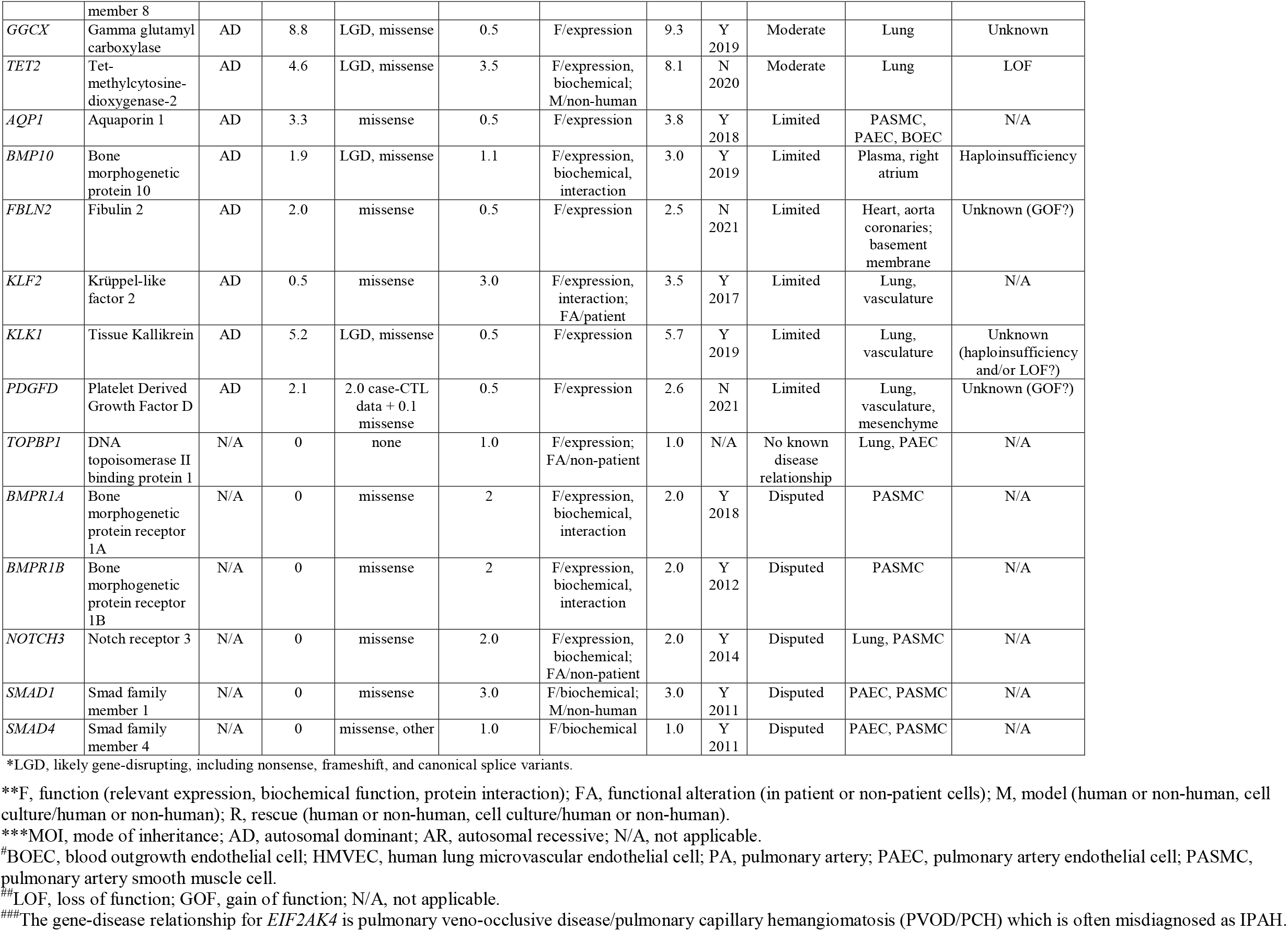
Strength of PAH-gene relationships for genes implicated in isolated H/IPAH.

**Figure 2.**
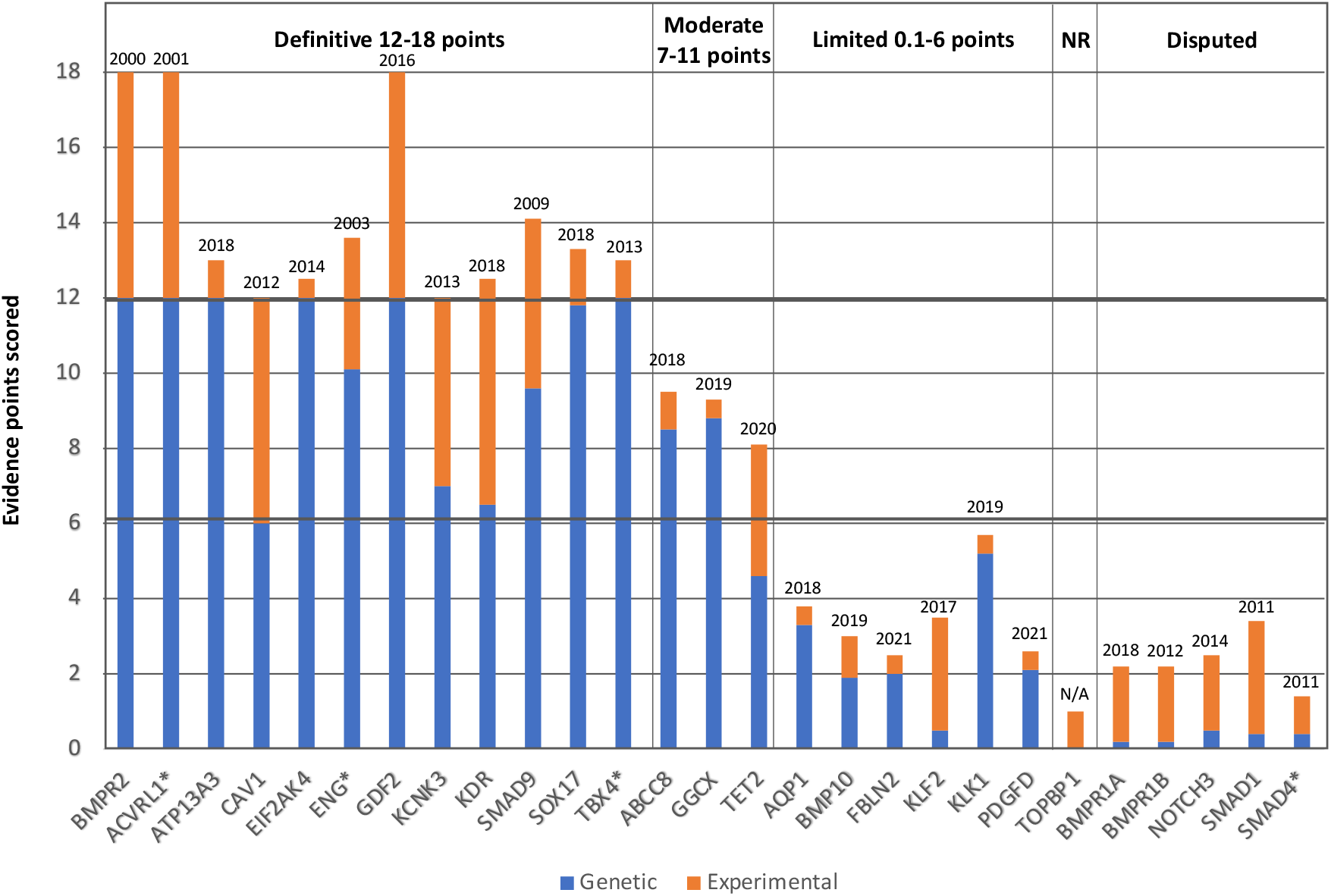
Quantitative contributions of genetic and experimental evidence to the clinical validity classifications of genes curated for PAH. The sums of genetic (blue) and experimental (orange) evidence scores are shown for genes classified as having definitive, moderate, or limited evidence of a monogenic relationship, no relationship (NR) or disputed relationship for H/IPAH or syndromic PAH*. Dates above the bars indicate date of first report of a gene variant identified in a PAH case.

#### Genes encoding members of the BMP pathway

##### Genes with definitive evidence

The discovery of a *BMPR2*-containing locus on chromosome 2 by linkage analysis in families with PAH was followed by identification of *BMPR2* LGD variants that segregated among affected family members (18, 19). Currently, more than 650 unique PAH-associated *BMPR2* variants have been reported (7, 10, 20). These include LGD and rare missense variants. The LGD variants comprise nonsense, frameshift, splice-site, and whole exon deletions, while the majority of missense variants localize to the conserved ligand-binding and protein kinase domains. Based on multiple large cohort studies, including the UK NIHR PAH cohort (7) and the PAH Biobank (10), we know that rare *BMPR2* variants cause approximately 70-80% of familial cases and 10-20% of sporadic cases, but are rarely found in cases with PAH associated with other diseases. *BMPR2* is expressed in pulmonary vascular cells, and reduced expression is observed in PAH patient-derived cells with and without *BMPR2* mutations (21, 22). BMPR2 is a type II receptor of the TGF-β superfamily that interacts with type I receptors BMPR1A, BMPR1B, and ALK1. These heterotetrameric complexes drive phosphorylation of SMAD molecules (SMAD1, SMAD5, SMAD8) to regulate transcription of nuclear target genes (23). In 2001, Morrell and colleagues (24), reported increased proliferation of pulmonary arterial endothelial cells (PAECs) derived from PAH patients with *BMPR2* truncating variants, demonstrating haploinsufficiency as a pathogenetic mechanism. These findings were confirmed in mouse and rat models heterozygous for *Bmpr2* null alleles (25-27) or transgenic for a dominant-negative mutation (28). In addition, the rodent models exhibited features of human PAH including increased right ventricular systolic pressure (RVSP) and increased arteriole muscularization. Rescue of RV function by wild-type BMPR2 (29) or BMPR2 ligand (30) provided definitive experimental evidence for a causal role of *BMPR2* mutations in PAH.

*CAV1* encodes caveolin-1, the main structural and signaling protein of caveolae. These specialized cell membrane lipid rafts play a key role in endocytosis, which is important for regulation of BMP receptors and downstream effectors, among others (31). Pathogenic variants in *CAV1* are rare causes of autosomal dominant and recessive lipodystrophies. *CAV1* was first identified as a causal PAH gene in 2012 by exome sequence analysis of four affected members of a multi-generational PAH family (32). A novel heterozygous frameshift variant in the final exon of *CAV1* co-segregated with disease but was also carried by several unaffected family members, indicating incomplete penetrance. Direct sequencing of 260 unrelated H/IPAH patients identified an additional *de novo* c.473delC (p.Pro158Hisfs*23) frameshift variant, located in the same region of the gene. Despite frameshift variants in the final exon being unlikely to trigger nonsense-mediated mRNA decay (33), immunostaining of lung sections from this IPAH patient revealed reduced CAV1 protein in small artery endothelial cells by comparison to healthy lung (32). Independent validation studies supported *CAV1* variants as a rare cause of H/IPAH, with seven additional rare variants reported to date (10, 13, 34, 35). CAV1 is expressed in lung endothelial and smooth muscle cells, and expression is decreased or absent in plexiform lesions (36). Investigation of patient fibroblasts expressing a *CAV1* c.474delA variant demonstrated reduced caveolae density and caveolar protein levels, and expression of the mutant protein caused reduced wild-type protein (37, 38). These data are consistent with a dominant negative mechanism of disease. Further, the increased SMAD1/5/8 phosphorylation observed in *CAV1* patient fibroblasts was not rescued by transduction with wild-type CAV1 (37), arguing against haploinsufficiency as the pathogenetic mechanism. Finally, *Cav1* knockout mice exhibit pulmonary vascular remodeling and features consistent with PAH, including higher pulmonary artery pressure, greater pulmonary vascular resistance, enhanced basal vascular permeability and RV hypertrophy (39, 40); and the phenotype is largely rescued by endothelial re-expression of *CAV1* (41). As the genetic variants associated with lipodystrophies are different from those associated with H/IPAH (with the exception of one reported pediatric case (42)) and the molecular mechanisms are likely to be different, *CAV1* was curated independently by our PH GCEP and the monogenic diabetes GCEP, and both curations are available on the ClinGen website.

*GDF2* encodes BMP9, a circulating ligand member of the BMP signaling pathway. Heterozygous variants in *GDF2* were identified as a potentially causal genetic risk factor in IPAH by burden testing analysis using the UK NIHR PAH cohort (7). Subsequently, two independent reports demonstrated evidence of *GDF2* variants in PAH (12, 43). Twenty-two discrete variants (6 LGD, 15 missense) were identified in the Han Chinese IPAH cohort alone (12). Taken together, these data identify a total of 32 predicted deleterious *GDF2* molecular defects including eight LGD variants. BMP9 ligand binding activates an endothelial specific BMPR-II/ALK1heterotetrameric complex to generate the intracellular phosphorylation relay routed through BMP-specific SMAD1/5/8 (44). *In vivo* analysis of BMP9 plasma levels comparing a *GDF2* variant-positive cohort (n=19) to unaffected controls (n=87) and IPAH patients without *GDF2* variants (n=38) confirmed a significant down-regulation of BMP9 in *GDF2* carriers. This finding was supported by the observation that secretion of BMP9 in cells transfected with *GDF2* variants was profoundly impacted. Treatment of PAECs with wild-type or mutant *GDF2* supernatant resulted in mutant-specific attenuation of the anti-apoptotic response (12). This initial gene curation indicated a strong gene-disease relationship between *GDF2* and PAH. Recuration three years after the initial gene discovery report (March 16^th^, 2022) identified four additional PAH patients with heterozygous variants, three of which were missense variants with *in vitro* evidence of loss of function, and one nonsense variant (45, 46). Furthermore, a combined analysis of the PAH Biobank and the UK NIHR PAH cohort identified *GDF2* as one of seven genes that were significantly associated with IPAH on a genome-wide basis (9).

The identification of homozygous *GDF2* LGD variants in two children with PAH raises the possibility of semi-dominant inheritance (47, 48). However, three other children under the age of ten years with homozygous truncating variants showed no evidence of PAH (confirmed by right heart catheterization in one) (49, 50), suggesting that homozygosity does not necessarily lead to early age of onset or more severe disease, and thus *GDF2* variants may act as a dominant gene in PAH susceptibility. There is also an emerging picture of overlap with HHT-like phenotypes, notably pulmonary arteriovenous malformations, that is beyond the scope of this manuscript (49-51).

The Mothers Against Decapentaplegic Homolog 9 (*SMAD9*) gene encodes SMAD8, involved in BMP signaling which tightly regulates processes related to development, differentiation, and growth (52). Shintani *et al*. (53) first associated a nonsense variant in *SMAD9* (c.606C>A, p.Cys202Ter) with PAH using a candidate gene screen of *ENG* and seven *SMAD*s in twenty-three Japanese IPAH patients without *BMPR2* or *ACVRL1* variants. A missense (54) and a unique nonsense variant (55) were then identified in two independent candidate gene screens. Fourteen additional variant carriers were identified by exome/genome sequencing of H/IPAH cases from the PAH Biobank (10), UK NIHR PAH cohort (7), and Han Chinese IPAH cohort (12). The variants included missense (n = 9, mostly located in receptor binding and phosphorylation domains), nonsense (n = 3) and an in-frame indel variant. *SMAD9* is ubiquitously expressed, including abundant expression in lung tissue (Genotype-Expression Project, GTEx, on May 1^st^, 2022) (56). SMAD8 was shown to undergo phosphorylation downstream of BMP type I receptors, inducing interaction with SMAD4 and transcriptional activity (57). SMAD8 p.Cys202Ter failed to immuno-precipitate with SMAD4 and transcriptional activity was diminished in a reporter assay (53). Transcript levels of *Id2*, a downstream target of BMP signaling, were reduced in patient PASMCs heterozygous for SMAD8 p.K43E, although response to ligand was largely preserved suggesting redundancy of SMAD1/5/8 function (54). *SMAD9* knockout mice are viable and show evidence of spontaneous pulmonary vascular remodeling in adult mice but other PH phenotypes have not been studied (58). A SMAD4-independent function of SMAD8 was identified in PAECs and pulmonary arterial smooth muscle cells (PASMCs) derived from lung explants of PAH patients. SMAD8-dependent post-transcriptional up-regulation of a subset of microRNAs was shown to exert anti-proliferative effects in control cells that was abrogated in cells from patients with *BMPR2* exon deletions or *SMAD9* p.R294X variant (55). The evidence supporting this gene-disease relationship has been replicated over more than 10 years without contradictory evidence.

##### Genes with limited evidence

In 2019, Eyries and colleagues identified *BMP10* as a new candidate PAH gene in the TGF-β/BMP family. Targeted sequencing of nine known PAH genes plus *BMP10* was performed in 263 patients, and a heterozygous nonsense variant and a missense variant in *BMP10* were found in two severely affected IPAH patients (59). Shortly afterwards, Gelinas and colleagues (43) performed exome sequencing of a pediatric cohort (n=18) and reported another missense variant in *BMP10*. Two more studies independently reported heterozygous missense variants in *BMP10* in two IPAH patients (60, 61). *BMP10* encodes the bone morphogenetic protein 10 (BMP10) ligand which binds to ALK1 in endothelial cells and activates the downstream phosphorylation of SMAD family transcription factors (62). BMP10 is a paralogue of BMP9 with 65% amino acid homology. *Bmp10* knockout mice die at an early embryonic stage due to retarded cardiac growth and chamber maturation (63). In contrast, *Bmp10* conditional knockout mice (induced postnatally) were reported to be viable and fertile, exhibiting no PH phenotype under normoxic or hypoxic conditions (64).

##### Disputed genes

*BMPR1A* and *BMPR1B* encode type I receptors integral to the canonical BMP signaling pathway. A relationship between *BMPR1B* and IPAH first emerged in a 2012 candidate gene study of 74 Japanese cases wherein two missense variants were described as pathogenic (65). However, based on a Japanese control cohort, 8.3KJPN (https://jmorp.megabank.tohoku.ac.jp/202109/variants), both variants were observed at a frequency exceeding the population prevalence of PAH. Additional publications reported variants in *BMPR1A* or *BMPR1B* associated with H/IPAH (10, 12, 34) but only two missense variants per gene met our inclusion criteria for potentially causal variants. Both BMPR1A and BMPR1B are expressed at equivalent levels to BMPR-II in human PASMCs but notably are indiscernible in PAECs (66). Both receptors are capable of binding BMPR-II with high affinity at the cell surface. Upon contact with extracellular ligand, a heterotetrameric complex of BMPR-II and either one of these receptors initiates a phosphorylation cascade via the SMAD pathway to regulate transcription of target nuclear genes (67). Due to a paucity of genetic evidence over time, both genes are disputed.

*SMAD1* and *SMAD4* encode TGF-β signal transduction proteins; receptor-regulated SMAD1 is BMP-specific whereas SMAD4 functions in the final pathway (downstream of BMPs/TGF-β/activins) with translocation to the nucleus and targeted gene transcription (68). Targeted sequencing of *SMAD1* and *SMAD4* in a cohort of 324 PAH cases led to identification of a *SMAD4* predicted splice-site variant, a *SMAD4* missense variant, and a *SMAD1* missense variant in three IPAH patients (54). Overexpression of the *SMAD1* variant resulted in reduced activity of a luciferase BMP-activated SMAD-binding reporter although its effect was modest compared to classical null *BMPR2* variation (54). Several other publications have since reported *SMAD1* and *SMAD4* variants in association with IPAH, but the overall number remains small (10, 12, 13). Both SMAD proteins are expressed in the human lung (GTEx on November 24^th^,2021) (56). SMAD1/4 protein levels were shown to be reduced in the rat monocrotaline model of PH (69) but there are contradictory reports in animal (70) and human lung tissue studies (71). *SMAD1* is a critical mediator of *BMPR2* signaling (72) with conditional deficiency – in endothelial or smooth muscle cells – predisposing mice to PH (73). However, human genetic data for both *SMAD1* and *SMAD4* remain weak.

#### Transporter and channel genes

##### Genes with definitive evidence

*ATP13A3* encodes a transmembrane cation transporter which was recently shown to transport polyamines (74). Polyamines are small metabolites required for normal cell growth and proliferation, and elevated plasma concentrations have been reported in multiple cancers and, more recently, PAH (75, 76). Monoallelic genetic variants in *ATP13A3* were first reported in the UK NIHR PAH cohort (6 LGD, 4 missense variants) (7). Independent validation of *ATP13A3* in PAH was demonstrated with the identification of four missense variants in the Han Chinese IPAH cohort (12) and five variants (2 frameshift, 1 nonsense, 2 missense) in H/IPAH cases from the PAH Biobank (10). More recently, biallelic *ATP13A3* variants were identified in three families with severe, very early onset PAH in five children with high mortality (77). *ATP13A3* is highly constrained for loss-of-function variants (pLoF = 1) (78) and most of the PAH-associated missense variants occur in conserved protein domains (79). Together, the data are consistent with a dose-dependent, semi-dominant mode of inheritance for *ATP13A3* variants. Functional evidence includes expression of ATP13A3 protein in PASMCs, PAECs, and blood outgrowth endothelial cells from IPAH patients (7) as well as decreased proliferation and increased apoptosis of blood outgrowth endothelial cells transfected with *ATP13A3* siRNA (7). Most of the protein-truncating variants are predicted to undergo nonsense-mediated decay indicating haploinsufficiency as the likely disease mechanism. For the missense variants, it is unclear whether the mechanism is loss or gain of function.

The *KCNK3* gene on chromosome 2p23.3 encodes a two-pore domain potassium channel, also known as TASK1. Heterozygous mutations in *KCNK3* were first reported by Ma *et al* in 2013 (80) as a cause of autosomal dominant PAH in a multi-generational family with exome sequence data. A novel missense variant was demonstrated to co-segregate with disease, but with incomplete penetrance. Targeted sequence analysis detected five additional missense variants in 5/320 unrelated H/IPAH cases, accounting for 1.9% of the total cohort. All variants were assessed by electrophysiological analyses, which indicated reduced current in mutant channels, supporting a loss of function disease mechanism (80). Similar findings have been reported for two variants identified in an independent Spanish cohort (81, 82). One of the variants was observed in a homozygous state in a consanguineous family (81), indicating potential semi-dominant inheritance for *KCNK3* variants. To date, more than 20 likely pathogenic missense variants have been reported in H/IPAH (7, 10, 12, 13, 34, 83-86). An additional family with early onset PAH and a homozygous 18 bp duplication was recently reported (87); however, inclusion of these data is pending re-curation of *KCNK3*. KCNK3 plays a role in the regulation of resting membrane potential and pulmonary vascular tone and is expressed in PASMCs (88). Moreover, both mRNA and protein expression are significantly reduced in lung tissue and isolated pulmonary arteries from PAH patients (89). PASMCs from IPAH patients (n=7) cultured with selective KCNK3 blocker A293 showed an 80% reduction of A293-sensitive current compared to controls (n=10), indicating that KCNK3 channel dysfunction is a key driver in the development of PAH (89). In mice, KCNK3 does not form a functional channel in PASMCs and is instead replaced by a KCNK6 channel (90, 91). *Kcnk3* mutant rats expressing a truncated channel demonstrate multiple features consistent with PAH, including age-related increases in RVSP, total pulmonary resistance, pulmonary vessel muscularization, and constriction of pulmonary arteries (92).

##### Genes with moderate evidence

*ATP binding cassette subfamily C member 8, ABCC8*, encodes sulfonylurea receptor-1 (SUR1), a regulatory subunit of adenosine triphosphate (ATP)-sensitive potassium channel, Kir6.2. Homozygous pathogenic variants in *ABCC8* are known to cause hyperinsulinemic hypoglycemia of infancy (93). In 2018, Bohnen and colleagues (94) first identified heterozygous pathogenic variants in *ABCC8* in twelve H/IPAH patients. Functional assessment of the variants by patch clamp and rubidium efflux assays demonstrated reduced channel function in comparison to wild-type in a cell culture model (94). Heterozygous missense and LGD variants were identified in more than twenty-one additional patients with H/IPAH (10, 14, 43). Identification of healthy heterozygotes in PAH families indicated incomplete penetrance (14, 94). Interestingly, rare predicted deleterious *ABCC8* variants have been identified in a similar number of patients with associated PAH forms (n = 19, not included in the curation for H/IPAH) (10, 95), suggesting a broader role for *ABCC8* in PAH. So far, only one child with PAH and hypoglycemia has been described with a heterozygous *ABCC8* nonsense variant (96). Based on differing modes of inheritanc and only a single patient exhibiting overlapping phenotypes, *ABCC8* was curated separately for PAH and monogenic diabetes. Expression of the SUR1 protein was confirmed in proximal pulmonary arteries and alveolar macrophages of IPAH patients (94). Further experimental evidence is required to elucidate the precise pathogenetic mechanism leading to PAH.

##### Genes with limited evidence

*AQP1* encodes the water channel aquaporin 1 protein, which not only facilitates water transport by supporting the osmotic water movement but also promotes endothelial cell migration and angiogenesis (97). *AQP1* was first implicated in pulmonary PAH in the UK NIHR PAH cohort (7). The study showed a significant enrichment of heterozygous predicted pathogenic variants in PAH patients compared to controls. Two missense variants were shown to co-segregate with the disease in three PAH families, with one healthy middle-aged variant carrier suggesting incomplete penetrance of *AQP1* variants (7). Wang and colleagues (12) identified three additional missense variants in the Han Chinese IPAH cohort. Expression of *AQP1* was demonstrated in lung endothelium of PAH patients and in human PAECs from healthy donors (7). In mice, homozygosity for *Aqp1* null alleles results in attenuated hypoxic PH (98), suggesting that complete LOF may not predispose to PAH. Thus, only heterozygous missense variants are currently considered to be potentially disease relevant. Due to the lack of further experimental data and the small number of PAH variant carriers, *AQP1* is currently classified as a gene with limited evidence for a relationship with PAH.

#### Growth and transcription/translation factor genes

##### Genes with definitive evidence

The Eukaryotic Translation Initiation Factor 2 Alpha Kinase 4 (*EIF2AK4*, also known as General Control Nonderepressible 2 (*GCN2*)) encodes a serine-threonine kinase present in all eukaryotes that can induce changes in gene expression in response to amino acid deprivation by binding uncharged transfer RNAs. Eyries *et al*. (99) and Best *et al*. (83) independently identified bi-allelic variants in this gene in patients diagnosed with PVOD (13 families) and PCH (1 family, 2 sporadic cases), respectively, suggestive of an autosomal recessive subtype of PAH. In subsequent years, further studies reported at least ten additional probands clinically diagnosed with PVOD/PCH and nine IPAH cases presenting at younger age and with reduced survival compared to patients without variants in *EIF2AK4* (100, 101). As with other recessive rare conditions, genetic diagnoses of *EIF2AK4* variants are more prevalent in consanguineous families. Experimentally, EIF2AK4 was detected by immunohistochemistry in lung tissue from an unaffected control and a PVOD patient without *EIF2AK4* variants, and was not detected in a PVOD patient with pathogenic variants in *EIF2AK4* (99).

The kinase insert domain receptor gene (*KDR*) is located on chromosome 4q12 and encodes the receptor for vascular endothelial growth factor type 2 (VEGFR2). Activation of VEGFR2 through ligand binding promotes cell proliferation, cell survival, and migration. LGD *KDR* variants in PAH patients were first reported in the UK NIHR PAH cohort (7). Subsequently, 2/311 PAH cases were identified with LGD *KDR* variants without detectable pathogenic variants in nine other PAH genes (102). The variants were associated with a low diffusing capacity for carbon monoxide (DLCO), and co-segregation analysis indicated that carriers of the variant either were affected by PAH or had decreased DLCO with the exception of one unaffected carrier (102). *KDR* is highly expressed in pulmonary endothelial cells (103). Mice exposed to hypoxia develop increased pulmonary arterial pressure, associated with minimal vascular remodelling (104). However, chronic hypoxia combined with SU5416-mediated inhibition of VEGFR resulted in vascular remodelling, PAEC proliferation and obliteration, and severe PH (105). Biomarker analysis revealed a signature like that observed in human PAH patients (105). Furthermore, endothelial-specific conditional deletion of *KDR* in mice resulted in a mild PAH phenotype under normoxia that worsened under hypoxia (106). Following a report of pathogenic *KDR* variants in four additional PAH patients (8) more than three years after the initial publication, the gene-disease relationship is classified as definitive for PAH.

*SOX17* is a two-exon gene encoding the SRY-box transcription factor 17, containing N-terminal DNA-binding HMG box and C-terminal β-catenin-binding domains. *SOX17* is critical in cardiovascular morphogenesis and postnatal vascular remodeling (107). A causal relation between heterozygous *SOX17* variants and PAH was first reported by gene burden testing in the UK NIHR PAH cohort (7). Subsequent studies have since supported the genetic association of *SOX17* with H/IPAH in independent populations including those of distinct genetic ancestry (10, 108-110). Together, these analyses identified 21 missense and predicted LOF variants in H/IPAH patients. In addition, data from a cohort of PAH associated with congenital heart disease patients (109) indicated that *SOX17* variants are especially enriched in this subclass of PAH with an early mean age-of-disease onset. The majority of nonsense and frameshift variants identified in PAH cases occur in the terminal exon, in the conserved β-catenin-binding domain; as such, the variant transcripts are predicted to escape nonsense-mediated decay and produce a dysfunctional protein. Wang *et al* (110), reported a terminal exon *SOX17* nonsense variant in a multi-generational family. *In vitro* luciferase assays indicated that the variant allele leads to a 14-fold reduction of target gene *NOTCH1* reporter activity and de-repression of β-catenin compared to wild-type. Intact NOTCH1 function is important for endothelial regeneration in response to injury, and endothelial-specific deletion of *Notch1* results in worsened PH in mice (111). Releasing the brakes on β-catenin expression promotes aberrant vascular remodeling. Immunolocalization of SOX17 clearly established endothelial specific expression in the pulmonary arterioles of wild-type cells and, importantly, also in patient vascular lesions (7).

##### Genes with limited evidence

*KLF2* encodes Krüppel-like factor 2 (lung KLF), a transcriptional repressor of inflammation, endothelial activation, and proliferation (112). In 2017, Eichstaedt and colleagues (113) screened an HPAH family with severe, rapidly progressive disease for copy number variants (*BMPR2, ACVRL1, ENG*) and exon/exon-intron boundary variants (31 known and candidate PAH genes). The affected father died at 32 years of age and two daughters were diagnosed at 25 years of age followed by lung transplantation. A heterozygous missense variant was identified in both affected daughters but not an unaffected brother. The variant is located at a hotspot for somatic lymphoma mutations, and functional analyses indicated loss of *KLF2* nuclear localization in patient-derived lung (114) and PAECs (115) as well as decreased transcriptional activity in transfected HEK T293 cells (114). No other *KLF2* variants have been reported in H/IPAH cases. KLF2 is highly expressed in human (116) and rodent (117) lung and decreased in PAH lung compared to healthy lung (115). Early overexpression studies showed proinflammatory gene expression activation and direct interaction with the endothelial nitric oxide synthase promoter in endothelial cells (118). Recently, decreased apoptosis and cell proliferation was demonstrated with *KLF2* overexpression in pulmonary vascular cells (115).

*PDGFD* encodes a member of the platelet-derived growth factor family, including four family members (PDGF A-D). PDGF-D is a mitogenic factor for cells of mesenchymal origin and is involved in regulation of embryonic development, cell proliferation, cell migration, survival and chemotaxis (119-121). *PDGFD* was recently identified as a PAH candidate gene in a combined analysis of the PAH Biobank and the UK NIHR PAH cohort (9). Rare variant burden testing was performed with exome/genome sequencing data from 1,647 unrelated European IPAH cases and ∼19,000 controls. In addition to five previously reported PAH genes, variant burden for two novel genes, *PDGFD* and *FBLN2* (see below), exceeded the Bonferroni-corrected threshold for significance. Nine IPAH cases were identified carrying seven unique *PDGFD* missense variants; two variants were recurrent in two unrelated cases each (9). Variants carried by at least five of the cases are predicted to disrupt a calcium-binding site or alter the conformation of a conserved PDGF/VEGF domain, but no functional data were provided. Gelinas *et al* (43) reported an additional IPAH case with a novel missense variant. At the experimental level, there is little direct evidence for a role of *PDGFD* in PAH. *PDGFD* is widely expressed, including lung (GTEx, on March 11^th^, 2022) (56) and arterial vasculature cells (122), but expression in pulmonary vasculature cell types has not been assessed. *Pdgfd* null mice exhibit decreased expression of heart development genes and increased systemic blood pressure but are otherwise phenotypically normal (123). Cardiac-specific *Pdgfd* transgenic mice have increased SMC proliferation, vessel wall thickening, fibrosis, heart failure, and premature death (124) but no data are available for pulmonary-specific overexpression.

##### No evidence for a PAH gene-disease relationship

*TOPBP1* encodes a binding protein that interacts with the C-terminal region of topoisomerase II beta. It is required for DNA replication, playing a role in the rescue of stalled replication forks and checkpoint control. TOPBP1 binds double-stranded DNA breaks and nicks, as well as single-stranded DNA. It also down-regulates E2F1 activity and inhibits E2F1-dependent apoptosis during G1/S transition and after DNA damage. *TOPBP1* was suggested as a possible IPAH locus following exome sequencing in twelve IPAH patients (125). However, the study focused on common variants (rs55633281, rs17301766 and rs10935070) with a minor allele frequency of greater than 5% in multiple populations. Subsequent analysis of more than 200 Italian IPAH patients showed no difference in the allele frequencies of these variants from the general European population or Tuscans from Italy, and no co-segregation between rs55633281 and PAH (126). Furthermore, no evidence for a role of rare *TOPBP1* variants was identified in the large UK NIHR PAH (7) or PAH Biobank (10) case-control studies, or the US/UK combined analysis (9). Functionally, *TOPBP1* expression was reduced in PAH lung tissues, especially in endothelial cells (125). In cultured PAECs, siRNA knockdown of control cells reduced *in vitro* tube formation on Matrigel and, conversely, over-expression of *TOPBP1* in IPAH cells partially rescued defective tube formation (125). Overall, data support that *TOPBP1* expression is altered in PAH lung tissues and it may play a role in PAH pathogenesis, but there is no genetic evidence for a gene-disease relationship.

##### Disputed genes

The Notch pathway is a highly conserved signalling cascade with important and diverse roles in human development and tissue homeostasis (127). Canonical Notch signalling is activated by ligand-receptor *trans*-interaction, leading to cleavage of the Notch intracellular domain, which complexes with nuclear co-activators to initiate the transcription of downstream genes, including the *HES* family. Chida *et al*. (128) used a targeted candidate gene approach including sequencing of *NOTCH3, HES1* and *HES5* in 41 IPAH patients with no identified variants in TGF-β/BMP family risk genes. The authors detected two *NOTCH3* missense variants; one variant, p.Gly840Glu, has a relatively high allele frequency in ethnically-matched controls (JPN8.3K, MAF: 0.0023) indicating that it is unlikely to be disease-causing. Furthermore, functional studies of the variant in HEK293 cell lines with tetracycline-inducible *NOTCH3* expression appeared contradictory (128). Although significant increases in proliferation and cell viability were observed, lower NOTCH3 levels were detected in mutant cells and luciferase reporter assays indicated impaired NOTCH3-HES5 signalling (128). Targeted sequencing in other cohorts (13, 85, 129-131) identified only two additional *NOTCH3* missense variants in H/IPAH that meet our curation thresholds. The NOTCH3 receptor is one of four Notch human homologues and regulates vascular homeostasis by maintaining smooth muscle cells in an undifferentiated state (132, 133). In the lung, NOTCH3 is predominantly expressed in small PASMCs and is well-documented to be important in the development of PAH; cultured PASMCs from IPAH patients display over-expression of both *NOTCH3* and *HES5* (134). Augmented NOTCH3 and HES5 levels also demonstrate a direct correlation with disease severity in hypertensive lung tissues from human and rodent models. *Notch3* knockout mice do not develop PH, supporting the paradigm that PASMC hyper-proliferation is driven by an up-regulation of NOTCH3 signalling through HES5 (134). However, the paucity of rare *NOTCH3* variants in PAH patients indicates that the regulation of NOTCH3 signaling is independent of genetic variation in the structural gene.

#### Other genes

##### Genes with moderate evidence

Gene burden testing in the PAH Biobank cohort (10) allowed the identification of two novel PAH candidate genes, gamma-glutamyl carboxylase (*GGCX*) and (tissue) kallikrein 1 (*KLK1*) (see below). Case-control analysis of 812 IPAH cases and 12,771 controls resulted in a variant burden for *BMPR2, GGCX*, and *KLK1* that exceeded the Bonferroni-corrected threshold for significance. *GGCX* encodes gamma-carboxylase, an enzyme essential for the activation of vitamin K-dependent proteins. *GGCX* plays important roles in blood coagulation, bone formation, vascular integrity, and inflammation (135). Biallelic variants in this gene have been reported with multiple phenotypes, amongst which vitamin K-dependent coagulation factor deficiency (MIM #277450) is best known (135). In the PAH Biobank, rare heterozygous *GGCX* variants (5 LGD, 9 missense) were identified in 18 H/IPAH cases (10). Three missense variants were recurrent in at least two cases each, and one missense and one LGD variant occurred in one IPAH and one PAH associated with another disease (10). Tissue-specific gene expression of *GGCX* has been detected in lungs and liver (3) but additional experimental data are required to elucidate a potential pathogenetic mechanism.

Tet-methylcytosine-dioxygenase-2, *TET2*, encodes an epigenetic regulatory enzyme involved in demethylation of cytosines. Somatic deactivating variants in *TET2* cause clonal hematopoiesis of indeterminate potential (136), a precursor to myeloproliferative diseases (137, 138). Somatic variants have also been implicated in cardiovascular disease (139, 140) and inflammation (141). To test the potential role of *TET2* in PAH, targeted rare variant burden analysis was carried out in the PAH Biobank cohort. Increased variant burden was demonstrated in PAH cases compared to controls, almost entirely due to LGD variants (9 LGD, 3 missense) and IPAH cases (8/12 cases) (11). Seventy-five percent were predicted germline and 25% predicted somatic. Hiraide *et al*. (142) reported a single germline variant identified in three Japanese H/IPAH patients but the minor AF (gnomAD all populations, East Asian, and Japanese) exceeded our threshold. In addition, the variant was curated as likely benign in ClinVar and an affected sister of an HPAH proband did not carry the variant. Anticipated increases in sequencing depth in the near future will likely increase variant identification among PAH cases for *TET2* and other clonal hematopoiesis of indeterminate potential genes. Experimental data included expression in lung (143) (GTEx, on March 23^rd^, 2022) (56), decreased circulating *TET2* expression in IPAH cases compared to CTLs (11), spontaneous PH in hematopoietic-specific mouse models (11), and increased circulating proinflammatory cytokines in cases vs controls and the mouse model (11). Interestingly, treatment of the mice with an IL-1beta inhibitor reversed the pro-inflammatory phenotype and PH (11).

###### Genes with limited evidence

Two PAH candidate genes with as yet limited evidence were recently identified in a combined analysis of the PAH Biobank and the UK NIHR PAH cohort (9). Rare variant burden testing was performed with exome/genome sequencing data from 1,647 unrelated European IPAH cases and ∼19,000 controls. In addition to five previously reported PAH genes, variant burden for *FBLN2* and *PDGFD* exceeded the Bonferroni-corrected threshold for significance. For *FBLN2*, a total of seven unique missense variants were identified in IPAH cases, including a recurrent c.2944G>T p.(Asp982Tyr) variant carried by four cases. This variant affects the last nucleotide of *FBLN2* exon 14 and is suggested to disrupt the splice donor site potentially leading to a null allele instead of a transcript with an amino acid substitution. *FBLN2* is located on chromosome 3p24-p25 and is a member of a family of eight fibulin genes. *FBLN2* is expressed in developing heart, smooth muscle precursor cells, developing cartilage, neural crest cells and endocardial cushion cells (120, 144, 145). Fibulin proteins are expressed and secreted as glycoproteins into the extracellular matrix, and have roles in developmental processes such as elastogenesis, embryonic organ development, tissue remodeling, and maintenance of the structural integrity of the basement membrane and elastic fibers. Fibulin-2 knockout mice are viable, fertile, and have intact elastic fiber formation. They exhibit attenuated angiotensin II-induced, TGF-β mediated, cardiac hypertrophy and myocardial fibrosis (146, 147) but have not been tested for PH.

*KLK1* is part of the kallikrein-kinin system that, together with the renin-angiotensin system, is involved in regulation of blood pressure. *KLK1* encodes a kininogenase contributing to the formation of the vasoactive peptide bradykinin. In the PAH Biobank (10), eight unique variants were identified (1 splicing, 1 frameshift, 1 nonsense, 5 missense), with three of the variants recurrent in at least two cases. *KLK1* is mainly expressed in the kidney, pancreas, salivary glands, brain, cardiovascular tissue and leukocytes. In addition, expression was reported in lung and vascular tissues (148-150). Overexpression of *KLK1* resulted in hypotension in transgenic mice whereas *KLK1* knock out mice were normotensive but showed blunted flow dependent vasodilatation (151-153). *KLK1* has been implicated in matrix degradation, smooth muscle cell migration, angiogenesis, and apoptosis - processes relevant to PAH (154-156) but PH has not been assessed.

### 2. Strength of evidence for genes implicated in syndromic forms of PAH

Three genes curated for syndromic forms of PAH (*ACVRL1, ENG, TBX4*), were all classified as having a definitive relationship with PAH (Table 2, Figure 2).

**Table 2.** Strength of PAH-gene relationships for genes implicated in syndromes including PAH.

| Gene | Gene name | Syndrome<br>* | Original<br>ClinGen<br>curation<br>GCEP | Original<br>classification | Genetic<br>evidence<br>for PAH | Variant<br>type<br>scored** | Experimental<br>evidence for<br>PAH | Evidence type<br>scored*** | Total<br>score<br>for<br>PAH | >3<br>yrs | Classification<br>for PAH |
| --- | --- | --- | --- | --- | --- | --- | --- | --- | --- | --- | --- |
| *ACVRL1* | Activin receptor<br>like 1 | HHT | Hemostasis/<br>thrombosis | Definitive | 12 | LGD,<br>missense | 6 | F/expression,<br>biochemical,<br>interaction;<br>M/non-human | 16 | Y<br>2001 | Definitive |
| *ENG* | Endoglin | HHT | Hemostasis/<br>thrombosis | Definitive | 10.1 | LGD,<br>missense,<br>other | 3.5 | F/expression,<br>interaction;<br>M/nonhuman | 13.6 | Y<br>2003 | Definitive |
| *TBX4* | T-box transcription<br>factor 4 | *TBX4*<br>syndrome | --- | --- | 12 | LGD | 1 | F/expression;<br>FA/patient,<br>non-patient | 13 | Y<br>2013 | Definitive |

\*HHT, hereditary hemorrhagic telangiectasia
\*\*LGD, likely gene-disrupting, including nonsense, frameshift, and canonical splice variants; CTL, control.
\*\*\*F, function (relevant expression, biochemical function, protein interaction); FA, functional alteration (in patient or non-patient cells); M, model (human or non-human, cell culture/human or non-human); R, rescue. (human or non-human, cell culture/human or non-human)

The activin A receptor like type 1 gene (*ACVRL1*), formerly known as activin-receptor-like kinase 1 (*ALK1*), is located on chromosome 12q13 and encodes the type I receptor, ALK1, involved in BMP signaling (157, 158). Variants in *ACVRL1* were first identified as causal for HHT, an autosomal dominant vasculopathy characterized by abnormal blood vessel formation in multiple organs (159). PAH was subsequently identified as a rare complication of HHT, with most cases attributable to *ACVRL1* (160-162). The variant types associated with HHT-PAH are mostly missense variants, located in the conserved protein kinase domain, but small deletions and nonsense variants have been identified in some cases. Within the protein kinase domain, Garamszegi *et al* (163) reported an enrichment of rare missense variants in a conserved nonactivating, nondown-regulating (NANDOR) box domain located in terminal exon 10 among HHT-PAH cases compared to HHT alone. The NANDOR box is required for downstream SMAD signaling (163); thus, variants associated with HHT-PAH that change one or more of the eleven amino acids defining the domain are likely pathogenic. Other rare HHT-PAH associated missense variants have been shown to cause subcellular mislocalization to the endoplasmic reticulum (161). *ACVRL1* is predominantly expressed in PAECs in normal arteries and PAH-associated plexiform lesions (160). In 2007, David and colleagues (44) identified BMP9 and BMP10 as the cognate ligands for ALK1 in endothelial cells. In the mouse model, homozygosity for a β-galactosidase-disrupted null allele demonstrated embryonic lethality with severe vascular malformations (164). Heterozygous mice developed adult-onset spontaneous PH with increased RVSP, RV hypertrophy, and vascular remodeling (165). Overall, genetic and experimental evidence are very strong and have been replicated by multiple groups over a twenty-year period.

*ENG* encodes endoglin, an accessory protein that interacts with ALK1 to promote TGF-β/BMP signaling (166, 167). Like *ACVRL1*, heterozygous LGD variants in *ENG* are predominantly associated with HHT (168). *ENG* variants were first associated with PAH in 2003, when two of eleven patients with HHT-PAH were identified with novel nonsense variants (161). Subsequently, Harrison *et al*. (169) reported a putative splicing branch site variant in intron 12 of *ENG* in a patient with HHT-PAH. Functional studies demonstrated resultant skipping of exon 13, leading to predicted loss of the *ENG* transmembrane domain. Since then, multiple other studies have identified *ENG* variants in HHT-PAH with an additional 4 nonsense and 5 missense variants, and an in-frame deletion (7, 10, 20, 170). *ENG* mutations were rarely reported in the absence of HHT, likely due to the much higher penetrance of HHT than PAH. However, in one case the diagnosis of PAH was made at three months of age, preceding the onset of HHT by eight years (169, 171). *ENG* is highly expressed in endothelial cells and plays an important role in angiogenesis (172, 173). Experimental evidence supporting a role for *ENG* in PAH comes from its function in TGF-β/BMP signaling and its interaction with ALK1 (166, 167). Endoglin is expressed in PAH and healthy lung endothelial cells (174). High expression observed in some PAH vascular lesions could be considered contradictory evidence; however, the analysis did not distinguish between L-endoglin and S-endoglin isoforms, which have opposing effects on TGF-β vs BMP signaling (175). Furthermore, *Eng* heterozygous knockout mice spontaneously develop PAH, characterized by increased RVSP, RV hypertrophy and reactive oxygen species levels (176).

T-box transcription factor 4 (*TBX4*) is a part of the T-box gene family located on chromosome 17q23.2 (177). *TBX4* is involved in early embryogenesis and plays a major role in lung branching morphogenesis and skeletal system development (178). Heterozygous LGD *TBX4* variants were first reported in families with small patella syndrome (SPS, OMIM #147891), an autosomal dominant skeletal dysplasia (179). A potential PAH locus at 17q22-q23.2 was first suggested by identification of a 3.75 Mb deletion that encompassed 15 known genes, including *TBX2* and *TBX4* (180). In 2013, Kerstjens-Frederiske *et al* reported two intragenic *TBX4* frameshift variants and one missense variant in patients with SPS and pediatric-onset PAH (181). Multiple other studies have since provided definitive genetic evidence that loss of *TBX4* function causes PAH, with numerous protein-truncating variants and missense variants that cluster in the T-box region of the gene (7, 10, 15, 34, 182). Notably, *TBX4* variants are more prevalent in pediatric PAH than adult-onset cases (34), and often associated with a *TBX4* syndrome involving PAH, other lung and cardiac anomalies, and SPS (183-185). Functional characterization of PAH-associated *TBX4* missense variants by an *in vitro* luciferase reporter assay demonstrated variant-specific LOF and gain of function effects (186). *TBX4* is strongly expressed in the lung during development (187), but experimental evidence for its role in PAH is currently limited. Cai *et al* showed phospho-SMAD1/5 levels were reduced in fetal lung fibroblasts following Crispr-Cas9 knockout of *TBX4*, and also in lymphocyte cell lines from two PAH patients with *TBX4* haploinsufficiency (188). SMAD phosphorylation is known to be attenuated in PAH, especially in cells with a *BMPR2* mutation. Overall, while the experimental evidence is somewhat limited, the genetic evidence is very strong and has been replicated by multiple groups over a nine-year period, leading to a definitive gene-disease relationship for *TBX4* and PAH.

## DISCUSSION

This report provides an overview of gene curation for twenty-seven genes implicated in H/IPAH using the semi-quantitative framework developed by ClinGen. Similar methodology has been applied to gene curation for at least six other cardiovascular diseases (189-194). To the best of our knowledge, this is the first study to evaluate gene-disease relationships in PAH using a standardized evidence framework. Based on available genetic and experimental evidence from peer-reviewed reports, twelve of the genes were classified as definitive, three as moderate, six as limited, one as having no known PAH gene-disease relationship, and five were disputed. These results offer up-to-date guidance to clinicians and genetic testing laboratories for molecular diagnosis and interpretation in isolated and syndromic causes of H/IPAH. Any future updates to the curations, including changes in clinical classification based on new evidence, can be tracked on the ClinGen website.

The role of the TGFβ/BMP pathway has been well-documented in the pathogenesis of PAH. While five genes in this pathway were classified as definitive, five genes were determined to have limited evidence. These include genes encoding BMP type 1 receptors (*BMPR1A, BMPR1B*) and SMAD signaling intermediaries (*SMAD1, SMAD4*), which were initially implicated through candidate gene analyses but have not been independently replicated over time. The *BMP10* gene was also classified as limited, potentially due to its relatively recent identification as a candidate causal gene; to date, only five variants have been reported in the literature. It should be noted that recuration over time may change the classification as new evidence is reported, which is particularly applicable for the newer genes with moderate to limited evidence.

With the recent expansion in the number of reported candidate genes, numerous diagnostic testing panels have been designed using high-throughput custom-capture sequencing. Although this method facilitates the screening of multiple risk genes at high coverage with reduced turnaround time, a major drawback is the need to regularly adapt the probe design with the rapidly changing genetic landscape, to incorporate newly identified genes. In addition, novel genes based on single reports are often included on panels to provide a more comprehensive analysis at reduced cost. However, our curation efforts have indicated that a number of these genes have limited evidence for causality, highlighting potential difficulties in variant interpretation. Based on our analyses, we therefore recommend a tiered approach, which prioritizes the analysis of strong and definitive genes first. For cases without a genetic diagnosis from the tier 1 panel, tier 2 and 3 analyses could then involve screening of moderate and limited evidence genes, respectively. Our investigations did not find any supporting evidence for *TOPBP1* or the set of five disputed genes, indicating that these genes should no longer be included on PAH routine genetic testing panels. Given the potential re-classification of limited evidence genes in the future, we encourage regular review of testing panels and adjustment of tiered analyses as appropriate. Most recently, the use of exome (or genome) sequencing for molecular diagnosis has become more cost-effective, providing coverage of most protein-coding genes without the requirement to adapt custom-capture probe sets. In addition, comparative analysis of sequence read depth between patient versus control samples may also be used to detect copy-number variation, providing greater flexibility for comprehensive molecular diagnosis than targeted genetic testing panels. Importantly, sequence data from undiagnosed cases can be stored and re-evaluated following evidence of new PAH gene-disease relationships.

Identification of a genetic cause of PAH in individual cases can have implications for clinical management including treatment (mono- vs multimodal therapy), surgical intervention and transplantation decisions, screening for associated conditions, and familial screening. A genetic diagnosis can lead to early treatment of associated medical conditions, cascade genetic testing of family members to identify those at risk for developing PAH, and clarification of reproductive risks to inform family planning decisions. For example, biallelic variants in *EIF2AK4* are diagnostic for PVOD/PCH (195), which can be difficult to diagnose clinically without a lung biopsy, and patients can be listed for transplant earlier in the course of disease which may improve outcomes. *ACVRL1/ENG* heterozygotes with PAH associated with HHT have a risk of arteriovenous malformations in brain, intestine, liver and lung (196); these patients require periodic MRI surveillance. *TBX4* variant carriers, especially children, have an increased risk of other lung, cardiac or skeletal defects (183, 186) and should be assessed by imaging studies and physical exam of the hands, hips, knees, and feet as PAH can overlap with SPS also caused by *TBX4*. Infrequent autosomal recessive forms of very early-onset, severe PAH have recently been identified for *ATP13A3* (77), *GDF2(47)* and *KCNK3(81)*. Such cases may be largely refractory to treatment and with high mortality, requiring early referral for surgery for a Potts shunt or lung transplantation (77, 197).

## Conclusions

Twelve genes have definitive evidence for causal effects of variants on PAH using a standardized evidence-based classification system and we will continue to regularly reassess the evidence for other genes.

## Data Availability

Gene curations are publicly available on the ClinGen website. Some of the curations are pending website upload (in progress).

https://search.clinicalgenome.org/kb/affiliate/10071

https://search.clinicalgenome.org/kb/affiliate/10028

## Nonstandard Abbreviations and Acronyms

APAH: pulmonary arterial hypertension associated with other diseases
ClinGen: Clinical Genome Resource
HPAH: heritable pulmonary arterial hypertension
IPAH: idiopathic pulmonary arterial hypertension
LGD: likely gene-disrupting
LOF: loss of function
PAECs: pulmonary arterial endothelial cells
PASMCs: pulmonary arterial smooth muscle cells
PAH: pulmonary arterial hypertension
RV: right ventricular hypertrophy
RVSP: right ventricular systolic pressure
SPS: small patella syndrome
VUS: variant of uncertain significance

## Author contributions

All named authors provided significant input to the conception and design of the study, data analysis and interpretation. All named authors were involved in preparation of the manuscript.

## PH VCEP members

Emily P. Callejo^1^, Kristina M. Day^2^, Daniela Macaya^3^, Gabriel Maldonado-Velez^2^

^1^Department of Pediatrics, Columbia University Irving Medical Center, New York, NY, USA.

^2^Indiana University School of Medicine, Indianapolis, IN, USA. ^3^GeneDx, Gaithersburg, MD, USA.

### PAH-ICON members

Stephen L. Archer^1^, Eric D. Austin^2^, Roberto Badagliacca^3^, Joan-Albert Barberà^4^, Catharina Belge^5^, Raymond L. Benza^6^, Harm Jan Bogaard^7^, Sébastien Bonnet^8,9^, Karin A. Boomars^10^, Olivier Boucherat^11,9^, Murali M. Chakinala^12^, Robin Condliffe^13,14^, Rachel Lynn Damico^15^, Marion Delcroix^16^, Ankit A. Desai^17^, Anna Doboszynska^18^, C. Greg Elliott^20^, Melanie Eyries^21,22^, Maria Pilar Escribano Subías^23,24,25,26^, Henning Gall^27^, Beatriz García-Aranda^23^, Stefano Ghio^28^, Ardeschir-Hossein Ghofrani^27,29^, Ekkehard Grünig^30^, Rizwan Hamid^31^, Paul M. Hassoun^15^, Anna R. Hemnes^32^, Katrin Hinderhofer^33^, Luke S. Howard^34^, Marc Humbert^35,36,37^, David G. Kiely^38^, Gabor Kovacs^39,40^, David Langleben^41^, Pablo Lapunzina ^42,43,44^, Allan Lawrie^13^, Jim E. Loyd^45^, Giovanna Manzi^46^, Jennifer M. Martin^47^, Evangelos D. Michelakis^48^, Shahin Moledina^49^, David Montani^36,50^, Nichols W. Morrell^51,52^, John H. Newman^32^, William C. Nichols^53^, Nuria Ochoa Parra^54,55^, Andrea Olschewski^39^, Horst Olschewski^39,40^, Dviya Pandya^47^, Silvia Papa^3^, Mike W. Pauciulo^53^, Roxane Paulin^8^, Roberto Poscia^3^, Steeve Provencher^11,9^, Rozenn Quarck^5^, Marlene Rabinovitch^56,57,58^, Laura Scelsi^28^, Werner Seeger^27^, Natascha Sommer^27^, Florent Soubrier^59^, Duncan J. Stewart^60^, Andrew Sweatt^61^, Emilia M. Swietlik^51^, Hemant K. Tiwari^62^, Roberto Torre^3^, Carmen Treacy^47^, Richard C. Trembath^63^, Olga Tura-Ceide^64,65,66^, Carmine Dario Vizza^3^, Anton Vonk Noordegraaf^67^, Martin R. Wilkins^34^, Roham T. Zamanian^68^, Dmitry Zateyshchikov^69^

^1^Department of Medicine, Queen’s University, Kingston, Ontario, Canada. ^2^Vanderbilt University Department of Pediatrics, Division of Allergy, Immunology, and Pulmonary Medicine, Nashville, TN, USA. ^3^Pulmonary Hypertension Unit, Department of Cardiovascular and Respiratory Sciences, Sapienza University of Rome, Italy. ^4^Department of Pulmonary Medicine, Hospital Clinic-IDIBAPS, University of Barcelona, Barcelona and Biomedical Research Networking Center on Respiratory Diseases (CIBERES), Spain. ^5^Laboratory of Respiratory Diseases & Thoracic Surgery (BREATHE), Department of Chronic Diseases & Metabolism (CHROMETA), Clinical Department of Respiratory Diseases, University Hospitals, University of Leuven, 3000 Leuven, Belgium. ^6^The Cardiovascular Institute, Allegheny General Hospital, Pittsburgh, PA, USA. ^7^Department of Lung Disease, Amsterdam UMC (location VUmc), Amsterdam, the Netherlands. ^8^Pulmonary Hypertension Research Group, Centre de Recherche de l’Institut de Cardiologie et de Pneumologie de Quebec, Quebec City, QC, Canada. ^9^Department of Medicine, Université Laval, Quebec City, Quebec, Canada. ^10^Department of Pulmonary Medicine, Erasmus MC, University Medical Center Rotterdam, the Netherlands. ^11^Pulmonary Hypertension and Vascular Biology Research Group, Institut Universitaire de Cardiologie et de Pneumologie de Québec, Department of Medicine, Université Laval, Quebec City, Quebec, Canada. ^12^Division of Pulmonary and Critical Care Medicine, Department of Medicine, Washington University School of Medicine, St. Louis, MO, USA. ^13^Department of Infection, Immunity & Cardiovascular Disease, University of Sheffield, UK. ^14^Royal Hallamshire Hospital, Sheffield, UK. ^15^Division of Pulmonary and Critical Care Medicine, Johns Hopkins University School of Medicine, Baltimore, MD, USA. ^16^Department of Pneumology, University Hospital Leuven, Leuven, Belgium. ^17^Indiana University, Indianapolis, IN, USA. ^18^Department of Pulmonology, Faculty of Medicine, University of Warmia and Mazury in Olsztyn, Olsztyn, Poland. ^19^Pulmonary Division, Intermountain Medical Center, Murray, UT, USA. ^20^Département de Génétique, AP-HP, Hôpital Pitié-Salpêtrière, Paris, France. ^21^INSERM UMRS 1166, Sorbonne Université and Institute for Cardiometabolism and Nutrition (ICAN), Paris, France. ^22^Department of Cardiology, Hospital Universitario 12 de Octubre, Madrid, Spain. ^23^Ciber-CV, Centro de investigación Biomédica en Red de Enfermedades Cardiovasculares, Madrid, Spain. ^24^Centro de Referencia Nacional de Hipertensión Pulmonar Compleja and ERN-Lung-Pulmonary Hypertension Referal Center, Madrid, Spain. ^25^Instituto de Investigación Sanitaria del Hospital Universitario 12 de Octubre (Imas12), Red SAMID, Madrid, Spain. ^26^Justus-Liebig University, Excellence Cluster Cardio-Pulmonary Institute (CPI) and Universities of Giessen and Marburg Lung Center (UGMLC), Member of the German Center for Lung Research (DZL), Giessen, Germany. ^27^Division of Cardiology, Fondazione IRCCS Policlinico San Matteo, Pavia, Italy. ^28^Department of Medicine, Imperial College London, London, UK. ^30^Center for Pulmonary Hypertension, Thoraxklinik Heidelberg gGmbH at Heidelberg University Hospital, Heidelberg, Germany and Translational Lung Research Center Heidelberg (TLRC), German Center for Lung Research (DZL), Heidelberg, Germany. ^31^Department of Pediatrics, Vanderbilt University School of Medicine, Nashville, TN, USA. ^32^Division of Allergy, Pulmonary and Critical Care Medicine, Vanderbilt University School of Medicine, Nashville, TN, USA. ^33^Laboratory of Molecular Genetic Diagnostics, Institute of Human Genetics, Heidelberg University, Heidelberg, Germany. ^34^National Heart and Lung Institute, Imperial College London, London, UK. ^35^Faculté de Médecine, Université Paris-Sud and Université Paris-Saclay, Le Kremlin-Bicêtre, France. ^36^INSERM UMR_S 999, Hôpital Marie Lannelongue, Le Plessis-Robinson, France. ^37^AP-HP, Service de Pneumologie, Centre de Référence de l’Hypertension Pulmonaire Sévère, Département Hospitalo-Universitaire (DHU) Thorax Innovation (TORINO), Hôpital de Bicêtre, Le Kremlin-Bicêtre, France. ^38^Sheffield Pulmonary Vascular Disease Unit, Royal Hallamshire Hospital, Sheffield, UK. ^39^Ludwig Boltzmann Institute for Lung Vascular Research, Graz, Austria. ^40^Medical University of Graz, Graz, Austria. ^41^Center for Pulmonary Vascular Disease, Cardiology Division, Jewish General Hospital and McGill University, Montreal, QC, Canada. ^42^Institute of Medical and Molecular Genetics (INGEMM)-IdiPAZ, Hospital Universitario La Paz-UAM, Madrid, Spain. ^43^CIBER Enfermedades Respiratorias, Centro de Investigación Biomédica en Red de Enfermedades Raras, ISCIII, Madrid, Spain. ^44^ITHACA, European Reference Network on Rare Congenital Malformations and Rare Intellectual Disability, Hospital Universitario La Paz, Madrid, Spain. ^45^Vanderbilt University Medical Center, Nashville, TN, USA. ^46^Department of Clinical, Anesthesiological and Cardiovascular Sciences, I School of Medicine, Sapienza University of Rome, Policlinico Umberto I, Rome, Italy. ^47^Department of Medicine, University of Cambridge, Cambridge Biomedical Campus, Cambridge, UK. ^48^Department of Medicine, Alberta Cardiovascular and Stroke Research Centre, University of Alberta, Edmonton, Canada. ^49^Great Ormond Street Hospital, London, UK. ^50^Université Paris-Saclay, AP-HP, French Referral Center for Pulmonary Hypertension, Pulmonary Department, Hôpital de Bicêtre, Le Kremlin-Bicêtre, France. ^51^Department of Medicine, University of Cambridge, UK. ^52^Department of Haematology, University of Cambridge, Cambridge, UK. ^53^Division of Human Genetics, Department of Pediatrics, Cincinnati Children’s Hospital Medical Center, University of Cincinnati College of Medicine, Cincinnati, OH, USA. ^54^Pulmonary Hypertension Unit, Department of Cardiology, Hospital Universitario Doce de Octubre, Madrid, Spain. ^55^Centro de Investigación Biomedica en Red en Enfermedades Cardiovasculares, Instituto de Salud Carlos III (CIBERCV), Madrid, Spain. ^56^Cardiovascular Institute, Dept of Pediatrics, Stanford University School of Medicine, Stanford, CA, USA. ^57^Division of Pulmonary and Critical Care Medicine, Dept of Medicine, Stanford University School of Medicine/ VA Palo Alto, Palo Alto, CA, USA. ^58^The Vera Moulton Wall Center for Pulmonary Vascular Disease, Stanford, CA, USA. ^59^Sorbonne Université, AP-HP, Département de Génétique, INSERM UMR_S1166, Sorbonne Université, Institute for Cardiometabolism and Nutrition (ICAN), Hôpital Pitié-Salpêtrière, Paris, France. ^60^Ottawa Hospital Research Institute, Sinclair Centre for Regenerative Medicine and the University of Ottawa, Ontario, Canada. ^61^Department of Pulmonary, Allergy and Critical Care Medicine, Stanford University, Stanford, CA, USA. ^62^Department of Biostatistics, University of Alabama at Birmingham, Birmingham, AL, USA. ^63^Department of Medical and Molecular Genetics, King’s College London, London, UK. ^64^Department of Pulmonary Medicine, Hospital Clínic-Institut d’Investigacions Biomèdiques August Pi I Sunyer (IDIBAPS), University of Barcelona, Spain. ^65^Biomedical Research Networking center on Respiratory diseases (CIBERES), 28029 Madrid, Spain. ^66^Department of Pulmonary Medicine, Dr. Josep Trueta University Hospital de Girona, Santa Caterina Hospital de Salt and the Girona Biomedical Research Institute (IDIBGI), Girona, Catalonia, Spain. ^67^Department of Pulmonology, Amsterdam Cardiovascular Sciences, Amsterdam UMC, VU University Medical Center, Amsterdam, the Netherlands. ^68^Department of Medicine, Stanford University Medical Center, Stanford, CA, USA. ^69^Federal Scientific Clinical Centre of Federal Medical and Biological Agency, Genetic Laboratory, Moscow, Russia.

